# Incidence, Persistence, and Steady-State Prevalence in Coding Intensity for Health Plan Payment

**DOI:** 10.1101/2025.04.18.25326080

**Authors:** Thomas G. McGuire, Oana M. Enache, Michael Chernew, J. Michael McWilliams, Tram Nham, Sherri Rose

**Author notes:** Correspondence to: Oana M. Enache, Encina Commons, 615 Crothers Way Stanford University, Stanford, CA 94305.

## Abstract

**Objective:** To define measures of Medicare diagnosis coding intensity that capture the dynamics of changes in coding practices.

**Study setting and design:** Retrospective analysis of coding for risk adjustment using observational claims data from Medicare beneficiaries.

**Data sources:** Enrollment and claims data from 2017-2018 of a random 20 percent sample of Medicare beneficiaries were subset to those assigned to an Accountable Care Organization in 2018.

**Principal findings:** We decompose prevalence of a diagnosis code into incidence (proportion of beneficiaries that newly have the code) and persistence (proportion of beneficiaries who previously had the code and continue to do so). Together these define steady-state prevalence, the hypothetical long-run prevalence implied by no changes in current rates of incidence and persistence of coding. Steady-state prevalence can help explain why observed prevalence tends to grow over time without continued behavioral change. For example, our measures suggest that the prevalence of the Specified Heart Arrhythmias diagnosis would continue to rise from 18.7% in 2018 to 28.0% without changes in coding practices.

**Conclusions:** Researchers and policymakers can better understand why changes in coding practices can take years to be fully reflected in data and monitor coding behavior by using our proposed measures.

**Callout Box:** *What is known on this topic:*

- Medicare incentivizes providers and insurers to code for as many diagnoses per beneficiary as possible, contributing to billions of dollars of Medicare program spending.
- Empirical research indicates that the frequency of diagnostic coding in Medicare has increased steadily in recent years.
- Current measures of coding practices for a given diagnosis largely count the prevalence, or the proportion of beneficiaries with a specified code, in one time period.

*What this study adds:*

- This study proposes decomposing coding practices over two years into several measures, which policymakers can use to better monitor coding behaviors and understand how they may change over time.
- This study defines steady-state prevalence as a novel measure of the ultimate prevalence of a code implied by current coding patterns hypothetically remaining unchanged.
- Applying our measures to Accountable Care Organization coding finds that observed coding prevalence will increase even without further behavioral change for the 10 diagnoses most impactful to 2018 risk adjustment.

## Introduction

The Centers for Medicare and Medicaid Services (CMS) finances care for Medicare beneficiaries using risk contracts in two major programs. The first is Medicare Advantage (MA), where healthcare plans are administered by contracted private insurers^1^. MA enrollment has grown rapidly in recent years and surpassed 51 percent of all eligible Medicare beneficiaries, or 31 million people^1^. Beneficiaries not enrolled in MA are said to be in Traditional Medicare (or TM) where their care might be financed through a second risk contract with a group of providers within the Accountable Care Organization (ACO) program^2^. Medicare assigns a beneficiary to an ACO largely based on their use of primary care services^3^.

Both MA and ACOs rely on the same baseline model, commonly referred to as the CMS-HCC model, to determine payment per beneficiary. In the name CMS-HCC and elsewhere, HCC refers to hierarchical condition categories, which roughly correspond to the diagnoses included as input variables in the model. CMS-HCC is a linear regression model with positive coefficients that predicts per-person spending as a function of each beneficiary’s diagnoses and demographic information^4^. For each enrollee, CMS obtains diagnosis codes from physician, hospital inpatient, and hospital outpatient claims from the previous year to predict costs for the performance year.

These risk scores are then used to adjust a benchmark for an ACO or MA plan’s beneficiaries according to their expected expenditures, an approach referred to as risk adjustment^4^. The MA and ACO programs risk adjust payments to MA plans and ACOs with the aim to provide adequate resources to care for beneficiaries and to incentivize insurers and provider groups to participate in the programs without avoiding high-cost patients^4,5^.

Although there are some differences in how payment is computed in ACO and MA contexts specifically, both of these risk-adjusted systems are premised on accurate information from providers and plans about patients’ health conditions. If existing coding is irregular or incomplete, then higher rates of coding by less intensive coders would improve the potential fit of risk adjustment payment formulas and the overall functioning of health plan markets. On the other hand, coding that does not accurately reflect a beneficiary’s health conditions can interfere with the efficiency and fairness of the risk adjustment payment formula. Both ACOs and MA plans are incentivized to code more intensely by the current payment system, as more diagnoses result in higher predicted spending by CMS-HCC^4,5^.

Prior research has established that coding intensity is substantially higher in MA than in TM^5–8^ and highly variable across MA plans and ACOs^9–11^. Higher coding intensity in the risk programs relative to TM for any reason increases federal spending on Medicare programs, making it important to understand trends in the prevalence of codes. This paper proposes and studies measures of coding intensity meant to capture the built-in dynamics associated with coding practices. In line with established terminology in epidemiology and other fields, we decompose the components of coding into incidence, the rate at which a code appears among those previously without the code corresponding to a given diagnosis, and persistence, or the rate at which a code continues among those with that code at an earlier time point.

To capture the long-run consequences of coded incidence and persistence, we also define steady-state prevalence, the ultimate prevalence of a code implied by the current rate of incidence and persistence if current coding patterns and underlying disease patterns in the population were to remain unchanged. This novel measure treats coding as analogous to a one-period Markov process. A positive gap between the steady-state prevalence implied by current patterns of coding and observed prevalence of a given HCC code signals that current coded prevalence will continue to increase even in the absence of any behavioral change in coding practices.

Persistence in coding has been studied previously^4,7, 12–14^, including one recent paper that used incidence and persistence to examine differential coding in MA compared to TM^7^. However, to our knowledge the dynamic implications of coded incidence and persistence have not been formally investigated to better understand coding behavior within a single Medicare program. In addition, steady-state prevalence is a novel measure and its use is a new approach to such analyses. To illustrate these concepts, we study incidence, persistence, and steady-state prevalence in Medicare’s ACO program with data from 2017 and 2018.

## Methods

This project was approved by the CMS privacy board and the Harvard Medical School Institutional Review Committee, which also waived the requirement for obtaining informed consent because all administrative data were deidentified.

### Data

We confined our study to the ACO sector in TM because the incentives for coding are stronger than in the non-ACO sector and the data were more readily available than MA data to researchers. We construct our measures from a random 20 percent sample of individuals continuously enrolled in Parts A and B of fee-for-service Medicare in both 2017 and 2018. We exclude beneficiaries who are either aged younger than 65 years, originally enrolled at least in part due to end-stage renal disease, or reside outside the United States. Our continuous enrollment requirement excludes beneficiaries who died or left the ACO program, implying that our measures are intended to be used for beneficiaries who are eligible to receive codes.

Our study sample consists of 1,799,068 beneficiaries who were assigned to an ACO in 2018 as identified based on the CMS ACO Shared Savings Program Beneficiary-level file. These exclusion and inclusion criteria produce a sample of 1,798,319 individuals. We used the 2017- 2018 version (version 22, or V22) of the CMS-HCC prospective risk adjustment formula to derive HCC variables from International Classification of Disease-10 codes. This version of the formula uses demographic information as well as 79 HCC variables that represent presence or absence of various diagnoses. The outcome variable of the V22 formula is projected healthcare costs per beneficiary in 2018.

### Statistical Analysis Procedure

One year of claims data contains the information needed to calculate *current coded prevalence,* 𝑝*_t_*, the proportion of beneficiaries with a disease code in a population in year 𝑡. Two years of data are required to measure coded *incidence,* 𝑖, and *persistence*, 𝑟, the rate of codes occurring among those previously without and with the code, respectively. 𝑝_t_ depends on 𝑝_t−1_ and the rates of coded incidence and persistence: 𝑝_t_ = (𝑝_t−1_)𝑟 + (1 − 𝑝_t−1_)𝑖. The *steady-state prevalence*, 𝑝, is a hypothetical future value of prevalence that will be reached if the current rates of coded incidence and persistence do not change. Our measure is based on an assumption that the underlying disease characteristics in the population remain unchanged. In a steady state, 𝑝 = 𝑝_t_ = 𝑝_t−1_ , so the steady-state prevalence satisfies 𝑝 = 𝑝𝑟 + (1 − 𝑝)𝑖, which simplifies to 𝑝 = 𝑖/(1 − 𝑟 + 𝑖). Therefore, steady-state prevalence depends only on incidence and persistence and can be computed directly from these two values independent of the current coded prevalence.

## Results

### Descriptive Data

Demographics of beneficiaries included as study participants are reported in Table 1, all of whom were enrolled in ACOs in 2017 and 2018. Geographically, 42.2 percent of participants reside in the South, 24.9 percent reside in the Midwest, 19.8 percent reside in the Northeast, and 13.0 percent reside in the Western United States. 9.2 percent of overall participants are eligible for Medicaid and 9.0 percent of overall participants have a disability. In terms of healthcare utilization, 17.0 percent of our sample had at least one inpatient stay and 73.7 percent of overall participants had at least one HCC code assigned in 2018, with each person having 2.1 HCCs recorded on average. Average annual spending per beneficiary overall is $7,911.10, in part because ACOs are not responsible for drug spending covered under Part D.

**Table 1.**
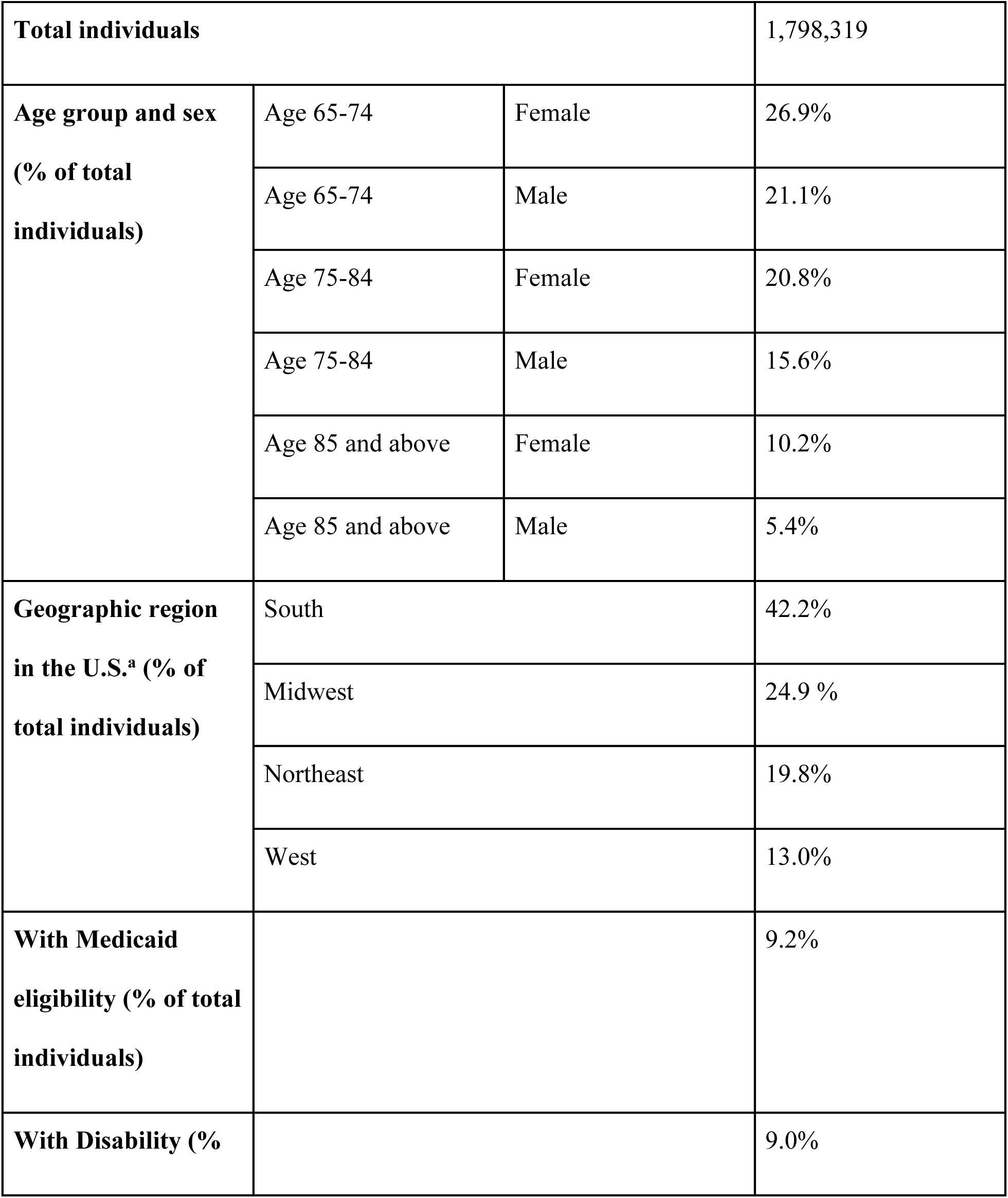
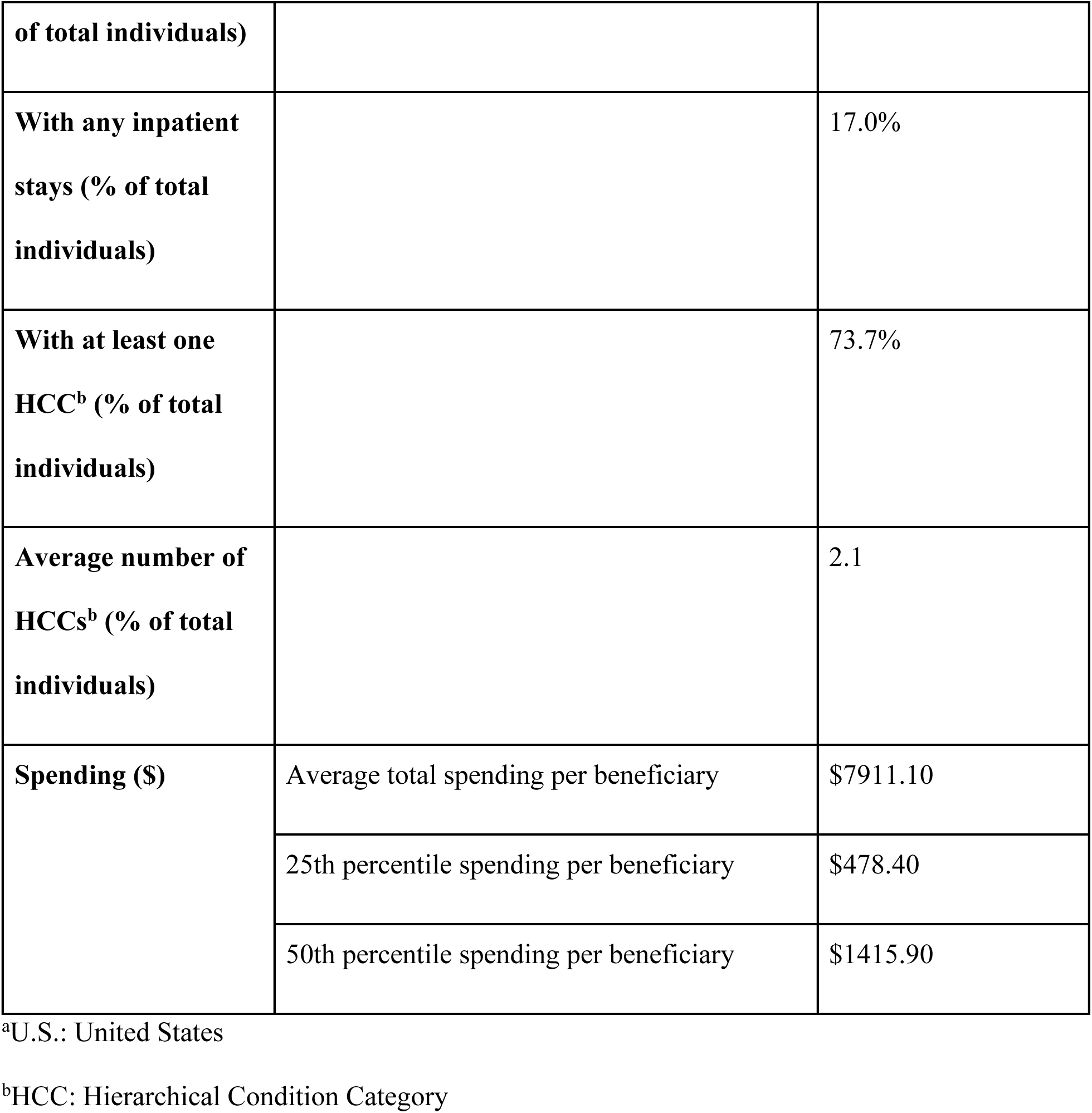
Demographic information for the analysis cohort, which corresponds to a random 20 percent sample of Medicare beneficiaries assigned to an Accountable Care Organization in 2018.

|  |  |  |  |
| --- | --- | --- | --- |
| **Total individuals** |  |  | 1,798,319 |
| **Age group and sex**<br>**(% of total**<br>**individuals)** | Age 65-74 | Female | 26.9% |
|  | Age 65-74 | Male | 21.1% |
|  | Age 75-84 | Female | 20.8% |
|  | Age 75-84 | Male | 15.6% |
|  | Age 85 and above | Female | 10.2% |
|  | Age 85 and above | Male | 5.4% |
| **Geographic region**<br>**in the U.S.<sup>a</sup> (% of**<br>**total individuals)** | South |  | 42.2% |
|  | Midwest |  | 24.9 % |
|  | Northeast |  | 19.8% |
|  | West |  | 13.0% |
| **With Medicaid**<br>**eligibility (% of total**<br>**individuals)** |  |  | 9.2% |
| **With Disability (%)** |  |  | 9.0% |

|  |  |  |
| --- | --- | --- |
| **of total individuals)** |  |  |
| **With any inpatient stays (% of total individuals)** |  | 17.0% |
| **With at least one HCC<sup>b</sup> (% of total individuals)** |  | 73.7% |
| **Average number of HCCs<sup>b</sup> (% of total individuals)** |  | 2.1 |
| **Spending (\$)** | Average total spending per beneficiary | \$7911.10 |
| | 25th percentile spending per beneficiary | \$478.40 |
| | 50th percentile spending per beneficiary | \$1415.90 |

<sup>a</sup>U.S.: United States
<sup>b</sup>HCC: Hierarchical Condition Category

### Measures

Table 2 depicts our measures for the 10 HCCs with the highest contribution to average risk scores among the ACOs in 2018, where contribution of an HCC to the risk score is the product of the 2018 current coded prevalence among ACOs times the weight of that HCC in the CMS-HCC community risk score formula. Together, these HCCs account for 39.8 percent of the average risk score. Incidence, persistence, current coded prevalence, and steady-state prevalence are computed using the formulas above. We illustrate some possible interpretations of our measures in these data below.

**Table 2.** Current coded prevalence, incidence, persistence, and steady state prevalence for the 10 hierarchical condition categories (HCCs) with highest risk score contributions in version 22 of the risk adjustment model for Accountable Care Organizations (ACOs) in 2018. HCCs are ordered by ACO risk score contribution.

| **HCC<sup>a</sup> Name** | **ACO<sup>b</sup> risk score contribution (%)** | **Incidence (%)** | **Persistence (%)** | **Current Coded Prevalence (%)** | **Steady-State Prevalence (%)** |
| --- | --- | --- | --- | --- | --- |
| Diabetes with Chronic Complications | 5.8% | 5.0% | 84.1% | 18.1% | 23.9% |
| Vascular Disease | 5.1% | 8.5% | 65.7% | 17.2% | 19.8% |
| Specified Heart Arrhythmias | 5.0% | 5.4% | 86.1% | 18.7% | 28.0% |
| Congestive Heart Failure | 4.6% | 6.2% | 72.6% | 14.2% | 18.4% |
| Chronic Obstructive Pulmonary Disease | 4.4% | 4.5% | 78.0% | 13.5% | 16.8% |
| Metastatic Cancer and Acute Leukemia | 4.2% | 0.8% | 70.9% | 1.6% | 2.7% |
| Major Depressive, Bipolar, and Paranoid Disorders | 3.5% | 4.1% | 70.1% | 8.8% | 12.0% |
| Rheumatoid Arthritis and Inflammatory Connective Tissue Disease | 3.3% | 3.3% | 66.9% | 7.9% | 9.0% |
| Acute Renal Failure | 2.3% | 4.3% | 30.1% | 5.3% | 5.7% |
| Morbid Obesity | 1.6% | 2.6% | 66.3% | 5.9% | 7.2% |

<sup>a</sup>HCC: Hierarchical Condition Category
<sup>b</sup>ACO: Accountable Care Organization

### Incidence

Coded incidence for diabetes with chronic complications, the HCC contributing the most to ACO plan payment, is 5 percent, meaning that among the enrollees without diabetes in a year, 5 percent are newly diagnosed as having diabetes compared to the prior year. Vascular disease, specified heart arrhythmias, and congestive heart failure all have incidence rates greater than 5 percent, which is perhaps not unexpected since about half of the population in our sample is 75 years or older (Table 1).

### Persistence

Table 2 also shows that most enrollees with one of the top ten HCCs continue to be coded with the HCC in a subsequent year (e.g., persistence is high). An exception is acute renal failure (currently known as acute kidney injury), which can improve with management and without dialysis for the majority of patients^15^ and is therefore unlikely to persist as a code over several years. However, rates of persistence, even for highly chronic conditions like diabetes and chronic obstructive pulmonary disease fall well below 100 percent.

Some unexpectedly low rates of persistence may be due to miscoding or undercoding. For example, an analysis of data from private health insurance in 2013 and 2014 found that persistence of the HCC indicating the very permanent diagnosis of paraplegia was only 57 percent^4^ and a more recent analysis of 2017-2019 data found that TM undercoding accounted for 22.3 percent of the difference in risk scores between TM and MA^14^. This suggests that such coding-related issues may be present across the insurance sector more broadly.

Lastly, the empirical distinction between current and steady-state prevalence can be illustrated by examining HCCs like HCC58 (Major Depression, Bipolar and Paranoid Disorders). These conditions are often, but not always, chronic and roughly half of patients with major depression experience just one episode in their lifetime^16^. Clinically, bipolar and paranoid disorders persist at higher rates, but even for these conditions, disease remission and recovery take place.

### Current and steady-state prevalence

In all cases, steady-state prevalence exceeds current coded prevalence, indicating that current coded prevalence is likely to increase over time for all ten HCCs even without changes in coding practices. In some cases, the gap between steady-state and current coded prevalence is particularly large, as in the 28.0 percent steady-state and 18.7 percent current coded prevalence for specified heart arrhythmias, signaling that particularly rapid increases in current coded prevalence can be expected for this condition.

We further illustrate this in Figure 1, which shows that current coded prevalence can continue to increase even if coding behaviors do not change for the same 10 HCCs. Incidence and persistence are held fixed at their 2017-2018 values. These are then used to calculate hypothetical projected prevalence values from 2018 current coded prevalence (time=0), which are plotted in subsequent years. Although the slope of each HCC’s projected prevalence varies over time, all of them increase for several years beyond 2018 and some (for instance, specified heart arrhythmias) increase for all ten years shown. This means that a one-time change in coding behavior alone could account for continued growth of ACOs or MA coding intensity (relative to non-ACO TM or TM coding respectively) for a significant amount of time.

**Figure 1.**
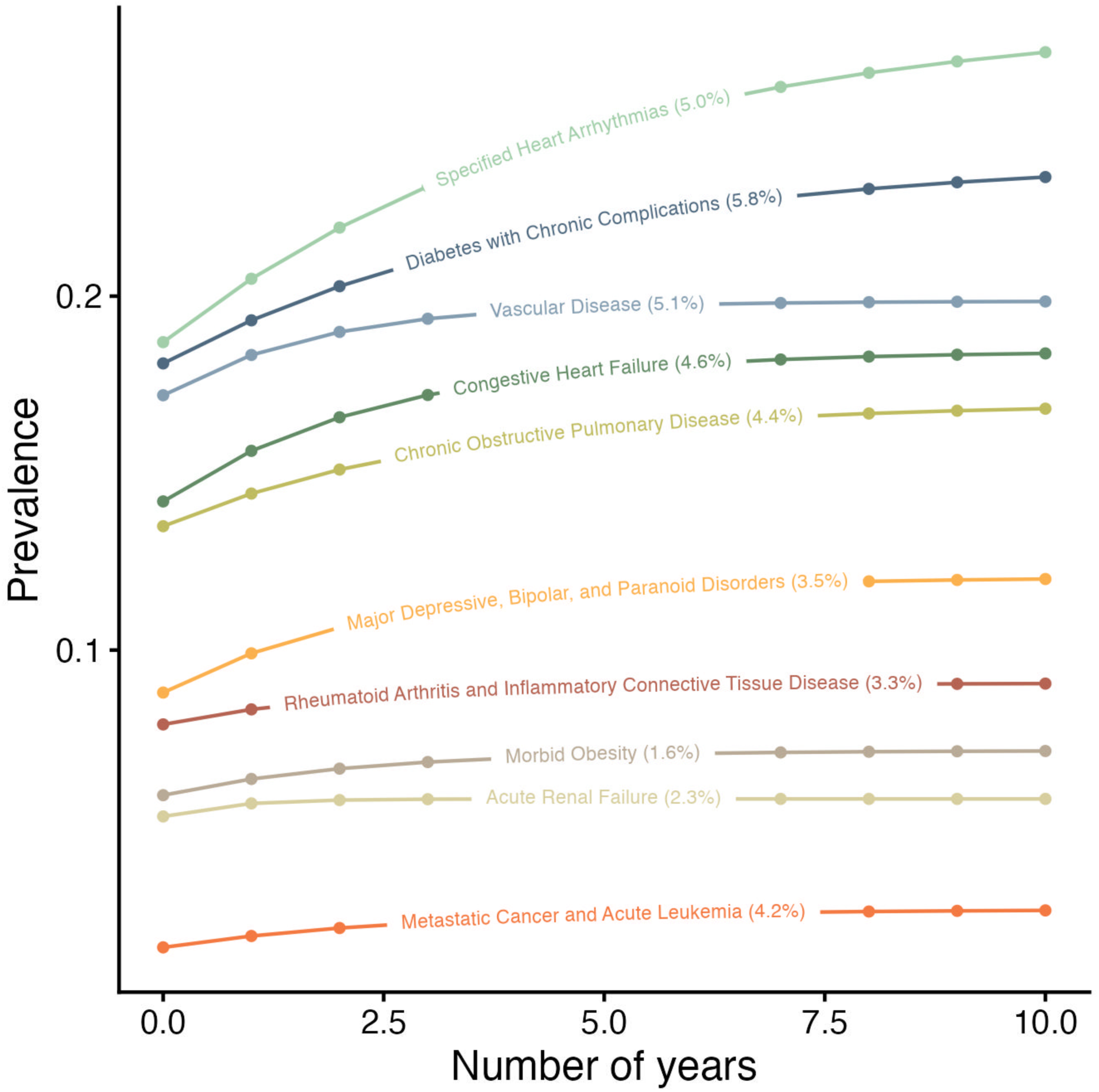
Current coding prevalence can continue to increase over time even if incidence and persistence do not change. Beginning at 2018 current coded prevalence values for coding in Accountable Care Organizations (ACOs) (shown on the y-axis at number of years = 0), each point within a given hierarchical condition category (HCC) represents the hypothetical current coded prevalence value if coded incidence and persistence for that HCC were to remain fixed as they were in 2017-2018. The percentage in parentheses following each HCC label corresponds to that HCC’s contribution to the ACO risk score. Even absent behavioral change (e.g., changes in coded incidence or prevalence), current coded prevalence can continue to increase for several years. The number of years can vary considerably; for instance, the HCC corresponding to acute kidney injury (labeled and formerly known as acute renal failure) has the same current coded prevalence after year 2, while the HCC corresponding to specified heart arrhythmias increases for all 10 years shown.

## Discussion

In most of the research and policy literature, the current coded prevalence of diagnostic codes is reported as the frequency of a code within a year’s data. Given disease chronicity, it is also useful to describe such coding as a pair of behaviors. Analogous to epidemiology, current coded prevalence in diagnostic coding depends on two underlying parameters: incidence and persistence of codes. Each of these behaviors are useful to monitor on their own and together define the steady-state prevalence implied by current coding practices.

Notably, viewing coding prevalence as a simple dynamic process has a number of limitations that can be improved upon in future research. First, our analysis is descriptive, not normative; our proposed measures do not by themselves answer the question whether changes in prevalence are due to better coding or opportunistic coding. Further, although we only consider two years of data for simplicity of computation, including more years in future analyses may be more informative. For example, a multiperiod Markov model or analyses involving more than two years of data would allow a richer characterization of the disease coding process but is not pursued here to keep the analysis straightforward. Doing so might be especially relevant in evaluating coding intensity changes as CMS-HCC model updates are phased in, which typically happens over the course of several years. Taking account of population trends due to entry and exit would modify our measures of incidence, persistence, and steady-state prevalence. We note that we illustrate our approach using an older version of the CMS-HCC model (V22 instead of the more current version) as it best aligns with our data; however, future work could extend to other model versions as well.

In addition, we do not explicitly define comparison populations to use in computing rates of coding; we do so intentionally to emphasize the flexibility of our proposed measures.

However, we note that additional comparison populations (for example non-ACO TM beneficiaries in comparison to ACO beneficiaries) may be informative to examine in assessing coding patterns. Although we only apply our dynamic characterization of coding in ACO populations, the measures we propose can readily be extended to other groups of individuals as well, including MA beneficiaries.

In any case, the novel measures we propose are a simple but informative tool for both clinicians and policymakers in a number of ways. Besides applications mentioned above, clinicians could evaluate the incidence and persistence of disease codes against the expected incidence and persistence of disease itself in a population, ideally using epidemiologic data free of coding incentives. Similarly, policymakers could compare incidence in codes from ACOs or MA plans to incidence from community-based surveys where indications of disease are not used for payment to inform further development in Medicare payment policies^11^. Steady-state prevalence can be used to better anticipate how coding might evolve even without behavioral changes. Lastly, our measures could also help policymakers better evaluate to what degree risk adjustment reforms (e.g., to dissuade upcoding) impact coding behaviors by distinguishing new coding after a given reform occurs from expected patterns.

## Conclusion

Concern about coding intensity plays a large role in payer policy and in the evaluation of the functioning of diagnostic-based risk adjustment systems, including but not limited to the CMS-HCC model. The decomposition of coding patterns into incidence and persistence, and the use of these to identify steady-state prevalence provide an informative tool to characterize coding and offer a partial explanation of why the observed prevalence of codes increases even in the absence of continued behavioral change.

## Data Availability

No data are available. Under the requisite data use agreements, data for the analyses are not publicly available.

## Acknowledgments

We thank Tim Layton, Richard Kronick, and members of the Risk Adjustment Network for comments on an earlier version. TGM, MC, JMM, and TN were funded by the National Institute on Aging of the National Institutes of Health under award number P01-AG032952. OME was funded by the National Library of Medicine grant 5T15LM007033- 40 and the Stanford Interdisciplinary Graduate Fellowship. SR was funded by the NIH Director’s New Innovator Award DP2-MD012722.

## References

1. Biniek JF, Freed M, Damico A, Neuman T. Half of All Eligible Medicare Beneficiaries Are Now Enrolled in Private Medicare Advantage Plans. Kaiser Family Foundation. Accessed June 15, 2023. https://www.kff.org/policy-watch/half-of-all-eligible-medicare-beneficiaries-are-now-enrolled-in-private-medicare-advantage-plans/

2. CMS Innovation Center Strategy Refresh. Centers for Medicare and Medicaid Services; 2021. Accessed April 10, 2024. https://innovation.cms.gov/strategic-direction-whitepaper

3. Assignment of Beneficiaries to Accountable Care Organizations Participating in the Medicare Shared Savings Program. Centers for Medicare and Medicaid Services; 2011. Accessed October 10, 2024. https://www.cms.gov/newsroom/fact-sheets/assignment-beneficiaries-accountable-care-organizations-participating-medicare-shared-savings

4. Ellis RP, Martins B, Rose S. Chapter 3 - Risk Adjustment for Health Plan Payment. In: McGuire TG, van Kleef RC, eds. Risk Adjustment, Risk Sharing and Premium Regulation in Health Insurance Markets. Academic Press; 2018:55–104. doi:10.1016/B978-0-12-811325-7.00003-8

5. Carlin CS, Feldman R, Jung J. The mechanics of risk adjustment and incentives for coding intensity in Medicare. Health Serv Res. 2024;59(3):e14272. doi:10.1111/1475-6773.14272

6. Medicare Payment Advisory Commission. Report to Congress: Medicare Payment Policy; 2024. Accessed April 30, 2024. https://www.medpac.gov/wp-content/uploads/2024/03/Mar24_MedPAC_Report_To_Congress_SEC.pdf

7. Kronick R, Chua FM, Krauss R, Johnson L, Waldo D. Insurer-level estimates of revenue from differential coding in Medicare Advantage. Ann Intern Med. Published online April 8, 2025. doi:10.7326/ANNALS-24-01345

8. Jacobs PD, Layton TJ. Identifying coding intensity in Medicare Advantage through switchers. Health Serv Res. Published online April 28, 2025:e14628. doi:10.1111/1475-6773.14628

9. Rose S, Zaslavsky AM, McWilliams JM. Variation In Accountable Care Organization Spending And Sensitivity To Risk Adjustment: Implications For Benchmarking. Health Aff . 2016;35(3):440–448. doi:10.1377/hlthaff.2015.1026

10. Chernew ME, Carichner J, Impreso J, et al. Coding-Driven Changes In Measured Risk In Accountable Care Organizations. Health Aff . 2021;40(12):1909–1917. doi:10.1377/hlthaff.2021.00361

11. McWilliams JM, Weinreb G, Landrum MB, Chernew ME. Use Of Patient Health Survey Data For Risk Adjustment To Limit Distortionary Coding Incentives In Medicare: Article examines use of patient health survey data for risk adjustment to limit distortionary coding incentives in Medicare. Health Aff (Millwood). 2025;44(1):48–57. doi:10.1377/hlthaff.2023.01351

12. Announcement of Calendar Year (CY) 2008 Medicare Advantage Capitation Rates and Payment Policies. Centers for Medicare and Medicaid Services; 2007. Accessed October 10, 2024. https://www.cms.gov/Medicare/Health-Plans/MedicareAdvtgSpecRateStats/Downloads/announcement2008.pdf

13. Copeland L. CMS Finalizes 2024 Medicare Advantage Payment Policies. Medicare Rights Center. April 6, 2023. Accessed October 10, 2024. https://www.medicarerights.org/medicare-watch/2023/04/06/cms-finalizes-2024-medicare-advantage-payment-policies

14. Datta NG, Chernew ME, McWilliams JM. Lack of Persistent Coding in Traditional Medicare May Widen The Risk-Score Gap With Medicare Advantage. Health Aff (Millwood). doi:10.1377/hlthaff.2024.00169

15. Vijayan A, Abdel-Rahman EM, Liu KD, et al. Recovery after Critical Illness and Acute Kidney Injury. Clin J Am Soc Nephrol. 2021;16(10):1601–1609. doi:10.2215/CJN.19601220

16. Goodwin FK, Jamison KR. Manic-Depressive Illness : Bipolar Disorders and Recurrent Depression (2nd Edition). Oxford University Press; 2007.

